# Multiple Sclerosis in the Digital Health Age: Challenges and Opportunities - A Systematic Review

**DOI:** 10.1101/2023.11.04.23298084

**Authors:** Bernhard Specht, Hana Jager, Samaher Garbaya, Alessandro Pincherle, Peiman Alipour Sarvari, Djamel Khadraoui, Reinhard Schneider, Ricardo Chavarriaga, Zied Tayeb

**Affiliations:** Myelin-H, 9 Av. des Hauts-Fourneaux, 4362 Esch-Belval Esch-sur-Alzette, Luxembourg & 71 Shelton St London, WC2H, United Kingdom; University of Luxembourg, 2 Av. de l’Universite, 4365 Esch-Belval Esch-sur-Alzette; Centre Hospitalier de Luxembourg, 4, rue Ernest Barblé L-1210 Luxembourg; Luxembourg Institute of Science and Technology, 5, Av. des Hauts-Fourneaux, 4362 Esch-Belval Esch-sur-Alzette, Luxembourg; Luxembourg Centre for Systems Biomedicine (LCSB), 7, avenue des Hauts- Fourneaux L-4362 Esch-sur-Alzette, Luxembourg; ZHAW Zurich University of Applied Sciences, Technikumstrasse 71, 8400 Winterthur, Switzerland; University of Lincoln, College of Health and Science, Brayford Pool, Lincoln LN6 7TS, United Kingdom

**Keywords:** Multiple Sclerosis, Digital Health, Remote Monitoring, Wearable Sensors, Personalised Medicine

## Abstract

In recent times, we have unequivocally witnessed a push towards digitising the healthcare system. Topics such as remote patient monitoring (RPM), digital health, and their use to monitor neurological disease progression have gained momentum and popularity. Notwithstanding the considerable advances that have been made in adopting such technologies and using them in the context of mental health or even a few neurodegenerative disease monitoring, they have not been widely used in the context of remote management and treatment of multiple sclerosis MS. In the same vein, given that (MS) is a very individualized disease to manage, there are numerous challenges yet opportunities associated with using digital health technologies for remote MS monitoring. This paper reviews the different research work and clinical attempts performed over the last decade (both home & hospital-based monitoring) en route to using digital health for MS monitoring and management. Similarly, this systematic review discusses the main challenges and barriers to translating that research from clinics into homes and highlights the opportunities in that context. Throughout this extensive review, we shine a light on various monitoring methods that hold the potential to be measured in a home environment, including electroencephalography (EEG) and evoked potentials (e.g., motor evoked potential (MEP), somatosensory evoked potentials (SSEP), and visual evoked potential (VEP)), electromyography (EMG), inertial measurement unit (IMU), and speech analysis. Combining such digital biomarkers could pave the way for developing a more personalised treatment for MS patients, thereby stopping its progression and avoiding silent MS disability. Adopting digital health for remote monitoring and management could also chart a route ahead for a new era of personalised medicine for MS patients and potentially other brain disorder patients.

## Introduction

More than 2.8 million people have MS, and the disease causes a reduction in life expectancy of 7–14 years^1^. Unlike any other brain disorder, MS is commonly diagnosed in the 20s & 30s and is the most common cause of disability in younger adults^2^. Although MS is incurable^3^, adequate treatment can manage symptoms, treat relapses, and slow the progression of MS. With more than 25 possible medications for MS (approved by the U.S. Food and Drug Administration (FDA))^4^, and a no-one-size fits all approach, it has become imperative to monitor medication effectiveness and, more importantly, monitor silent disease progression and any cognitive decline in MS patients. In that context, MS monitoring and management could greatly benefit from the recent advances in digital health and RPM. Notwithstanding the plethora of systematic reviews of digital health and wearable sensors for monitoring and managing other neurodegenerative diseases, such as Parkinson’s or dementia, fewer systematic reviews have covered digital health and RPM methods in MS management and monitoring. Along the same lines, these few review studies tend to have a narrow focus by examining studies covering one single aspect of the disease, such as assessing one particular symptom, reviewing previous studies covering one single modality (biosignal)^5^, or they tend to focus solely on mobile apps using questionnaires^6^, excluding, therefore, the plethora of previously conducted clinical studies whose focus spans from monitoring the cognitive decline in MS patients to assessing & monitoring other symptoms such as speech difficulties, muscle fatigue, balance & gait problems, cognitive decline and demyelination, using EEG, speech, EMG, IMU data, and other wearable data. Over and above, throughout our literature review, it is worth noting that discussing the challenges and barriers to translating research from the various clinical studies into homes and highlighting the opportunities is often an overlooked question and a missing component. Unlike previous review studies, this systematic review of the literature is intended to be more comprehensive. It, therefore, includes various monitoring methods that hold the potential to be measured in a home environment, namely EEG, EMG, speech, digital apps, IMU, as well as other wearable data. Hence, this review paper should serve as a comprehensive overview of the previously performed clinical research work on home & hospital-based MS monitoring performed over the last decade. Consequently, this review’s findings and conclusion should enhance the reader’s understanding of the gaps in the literature and the obstacles to fully embracing the use of digital health technologies and wearable sensors to monitor MS progression and its treatment effectiveness, which could contribute to stopping its progression and avoiding disability. The remainder of this paper is structured as follows: the first section describes our search strategy and studies selection. The second section is split into different sub-sections covering and summarising previous clinical work on EEG and evoked potentials, IMU & motion capture, EMG, digital apps, speech, and other wearable data. The last section enumerates the strengths, weaknesses, and existing gaps of the reviewed clinical studies and discusses veiled opportunities, as well as possible future improvements in MS monitoring and management using digital health technologies.

## Material and Methods

### Literature Search Criteria & Strategy

The PubMed database was used to evaluate and review previous research work and clinical attempts for both home & hospital-based monitoring of MS. The search strategy involved combining a match of “multiple sclerosis” in either the title or abstract field with a match of the following search terms in the abstract or title: “wearable”, “digital health”, “mhealth”, “sensor”, “EEG”, “electroencephalography”, “evoked potentials”, “EMG”, “electromyography”, and “speech”. Furthermore, abstracts of the identified references were carefully examined to exclude those deemed irrelevant to the review. The search was limited to the English language and reviewed papers published from January in the last decade.

### Study Selection

The following criteria were applied to filter and identify eligible studies:

1. Objective Measures Provided by Sensors: Studies without objective measures provided by sensors were excluded, as they did not align with the focus of this review on digital health technologies for monitoring multiple sclerosis (MS).
2. Sample Size: Case studies and studies with fewer than 10 MS patients were excluded from prioritizing studies with larger sample sizes that provide more robust insights.
3. Mixed Conditions: Studies focusing on mixed conditions or comorbidities alongside MS were excluded to maintain a specific focus on MS monitoring.
4. Pediatric MS: Studies explicitly focusing on pediatric MS were excluded
5. Reviews: Review articles were excluded from the selection process to ensure a primary emphasis on original research studies.
6. Specific Subgroups: Studies focusing solely on specific subgroups of MS patients (e.g., only those with sleep problems) were excluded to maintain a broader perspective on MS monitoring.
7. Language: Only studies available in English were considered
8. Abstract Availability: Only studies with available abstracts were considered
9. Timeframe: Only studies published within the last ten years were included to incorporate recent advancements and developments in the field.

In the initial identification phase, we queried the PubMed database, resulting in the identification of a total of 756 papers. Subsequently, in the screening phase, we scrutinized the abstracts of these papers and narrowed down the selection to 107 papers, each of which exhibited substantial relevance to our study. These 107 papers were categorized into various subdomains, including 12 on speech, 61 on EEG and evoked potentials, 12 on IMU, 14 on EMG, 4 on digital apps, and 5 addressing other pertinent aspects.

## Results

This section provides an extensive literature review of previous clinical attempts to use sensor-based data to monitor MS. We primarily focus on EEG, EMG, speech, IMU, and digital apps, given the potential they hold to be used in a home environment. An overview of the different reviewed biosignals is depicted in Figure 1.

**Figure 1.**
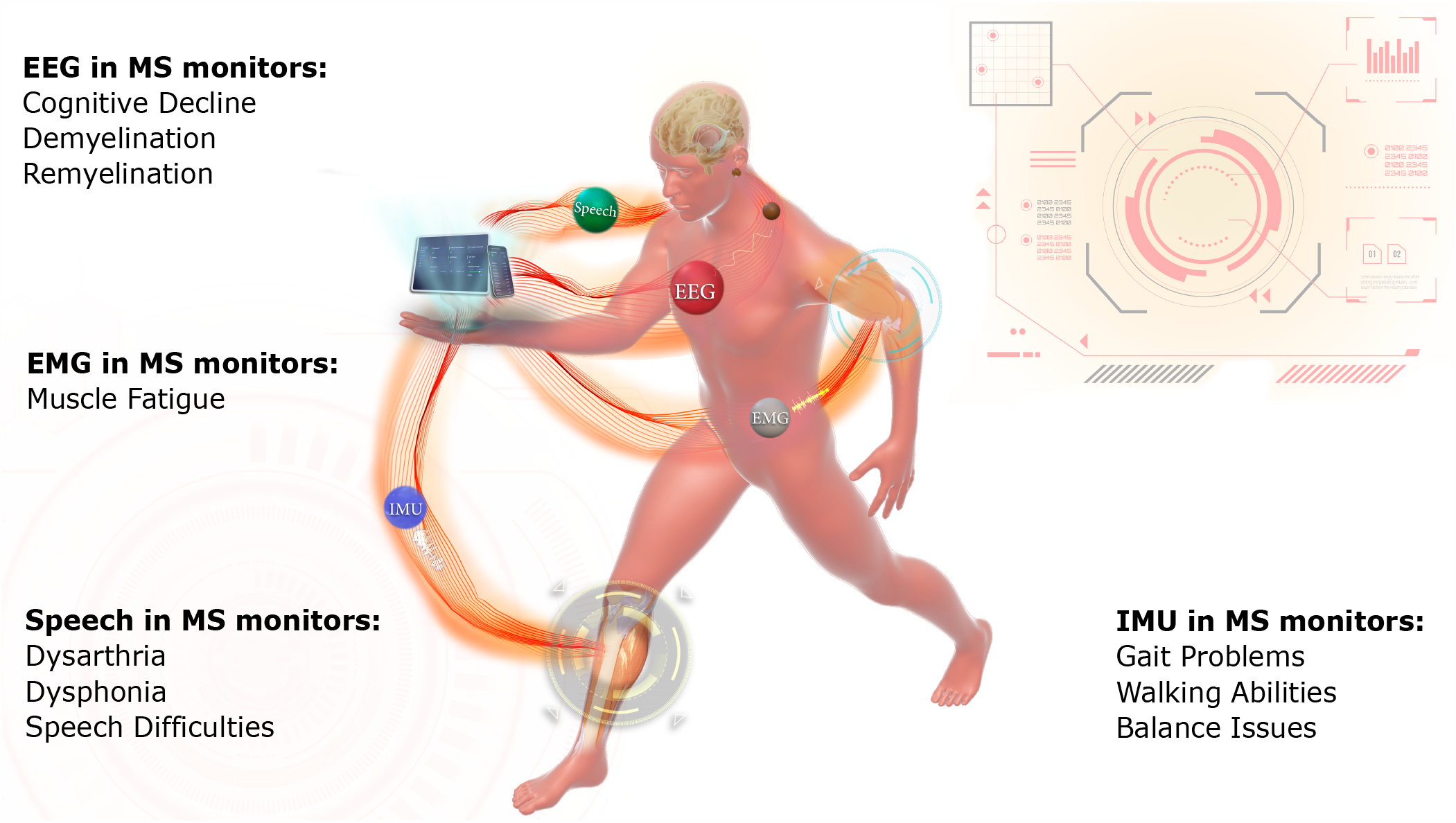
An overview of the key biosignals essential for remote MS monitoring, along with the possible digital biomarkers that could be derived from these signals in the context of MS home-based monitoring.

### EEG and Evoked Potentials

Throughout our literature review, the vast majority of the previous EEG studies and research work were on VEP, and focused on analysing its latency and amplitude^7–37^ for monitoring and diagnostic purposes. In the same vein, the second most frequently investigated evoked potential in EEG is MEP, whose latency, amplitude and any detected abnormalities have extensively been analysed and investigated^14, 20, 21, 25, 31, 32, 38–45^. Interestingly, SSEPs^10, 17, 20, 21, 28, 35, 36, 43, 46–48^ and AEPs^8, 16, 17, 28^ were less common and frequently used in the reviewed studies, but still offer, when combined with other evoked potentials, solid composite scores. Similarly, authors in^49, 50^ investigated and analysed evoked related potential (ERP) responses in MS patients by using the two-tone oddball paradigm. In a group of 16 MS patients and 19 healthy controls, Paolicelli et al.^49^ observed a latency when detecting the P300 peak in MS patients compared to healthy controls. They revealed an inverse correlation between the P300 amplitude and the fatigue level. Furthermore, evoked potentials in EEG have been analyzed during resting state^51–60^, during attention tasks^61–64^ or motor tasks^42^. Authors in^11^ showed that a combination of evoked potentials (EPs) could predict MS progression in terms of Expanded Disability Status Scale (EDSS) after 20 years. In another study, authors in^47^ observed that tongue somatosensory evoked potentials (SEPs) could reveal significant worsening of the trigeminal sensory pathway over four years. Similarly, Chinnadurai et al.^46^ highlighted that extracted latencies from VEP and SEP could serve as accurate predictors of falls in MS patients within one year. Along the same lines, authors in^20, 21, 25, 28, 31^ postulate that combined scores are a good predictor for EDSS in MS patients longitudinally. The detection of VEP from EEG signals was investigated, and the quantification of its amplitude and latencies between different MS patients were studied in^7, 15, 18, 23, 27, 37^. For that, authors in^7, 15, 18, 23, 27, 37^ postulate that patient with multiple sclerosis (PwMS) have in average longer VEP latencies when their retinal fibres are below norm. When analysing resting state in EEG, authors in this study^58^ unravelled that PwMS showed an increase in theta and a decrease in alpha (relative and absolute). Similarly, Babiloni et al.^55^ found that alpha and delta activities in the resting state were lower in PwMS. Conversely, an increase in alpha and gamma was found in^57^. Keune et al. showed a correlation between increased alpha 1 and 2 activity, as well as increased frontal theta*\*beta ratio with worse performance in the symbol digit modalities test (SDMT) test for MS^53^. High theta*\*beta ratio correlated with slow SDMT speed^60^. Along the same lines, when analysing resting state EEG, authors in^65^ observed that PwMS have significantly lower posterior dominant rhythm and amplitude, which was negatively associated with disability. When analysing the correlation between evoked potentials in EEG and MS progression, authors in^48^ revealed that SEP in EEG has a more pronounced correlation with MS disability, i.e. EDSS worsening, compared to MRI scans. Likewise, a combined score of EPs showed a stronger correlation with baseline EDSS than MRI white lesion burden^28^, pinpointing, therefore, the potential of evoked potentials in EEG in monitoring and predicting silent disability progression. Overall, it was shown that lower-limb MEPs correlated with worse walking performance, whereas upper-limb MEPs correlated with worse dexterity as measured by 9-hole peg test (9HPT). Interestingly, Gschwind et al.^52^ observed a pronounced correlation between resting state EEG and important MS monitoring parameters, namely annual relapse rate, disability score, depression score and level of fatigue. Along the same lines, authors in^63^ postulate that ERPs during the visual oddball task are correlated with processing speed at baseline in PwMS. Moreover, brain stem reflexes show correlations with clinical, EP and MRI findings and, combined with EPs, effectively predict brain stem dysfunction^17^. Reduced interhemispheric connectivity correlates with changes in visual ERPs in patients with relapsing-remitting MS (RRMS). PwMS also showed increased corticospinal excitability while observing suboptimal 9HPT performances on video^45^. When investigating fatigue (mental and physical) in PwMS, a previous study^49^ showed that P300 amplitude is inversely correlated with fatigue with an increased latency when PwMS were compared with HC. Fatigue, assessed by the modified fatigue impact scale (MFIS) score, was, however, correlated with an increased small world index (a measure for the efficiency of information transfer) in the sensory network. Global field power in EEG bands decreased for healthy control (HC) after blocks of muscle contractions, while PwMS showed no significant changes^66^. In a different clinical application, VEPs in EEG have been widely used and investigated as an indicator for treatment responsiveness^34^ before. While on treatment with Clemastine, patients showed a reduction of 3.2ms in the P100 component, which after treatment still was 1.7ms above the norm^9^, albeit the study was only conducted with 25 patients. In the same vein, combined EP scores improved after Fingolimod therapy^20^. Plasmapheresis and immunoadsorption in patients with steroid-refractory relapses showed a significant improvement in VEP latencies^24^. While scores, assessed by PASAT and BDI improved after Dalfampridine treatment, VEP remained the same^30^. High doses of biotin made P100 reappear and normalized latencies in a few patients^33^. MEP measures and 9HPT improved after treatment with Ocrelizumab^43^. After steroid treatment, rapid changes in the excitability of the motor cortex can be observed^44^. EDSS did not reflect any changes after treatment with interferon beta and Glatiramer, but VEP scores improved in the Interferon beta group^35^. It is worth noting that other studies and treatments show no significant changes in electrophysiological changes^10, 12, 32^.

### IMU and Motion Capture

When conducting this extensive review, we found out that the vast majority of the studies used walking tasks to collect and analyse IMU and other motion-capture-based data^67–73^ to assess gait and balance issues in PwMS. Overall, motion capture devices have revealed significant correlations between gait indices and subjective and objective walking abilities. Such indices include normalized velocity, stride length and hip range motion^67^. PwMS demonstrated a higher variability in their lumbar yaw range and lateral food deviation, particularly after turning and stepping over an object^68^. Similar to this finding, another study found more asymmetrical sway patterns when comparing PwMS with HC^69^. Levels of disability and self-reported fatigue in PwMS were found to correlate with changes in gait parameters like step and stride regularity and swing time asymmetry^70^. Differences in stride length and swing time between PwMS and HC have also been identified by Bourke et al.^73^. Aside from that, gait parameters have also shown reliability and reproducibility in between-centre studies when using different equipment^74^. A few gait bouts (6 cycles) can be sufficient for providing stable features^71^ when assessing and monitoring PwMS. When shining some light on the use of IMU and motion capture data for rehabilitation, previous studies^72^ demonstrated that gait parameters, such as cadence, velocity, stability and balance, can be used to rate the effect of neurorehabilitation. Furthermore, authors in^75, 76^ showed that accelerometer-based performance metrics revealed instabilities and were able to separate groups of fallers and non-fallers after 30 seconds of sit-stand transitions. Similarly, akin studies have shown a correlation between accelerometer-based performance metrics and MS disease severity, fatigue and balance confidence^77^. While suggested as suitable for remote MS monitoring^77^, measures can be different in unsupervised settings^75^. Over and above, authors in^78^ highlighted that PwMS showed a reduction in sway complexity paired with an increase in sway intensity while performing an altered Romberg test (patient stands on foam), which could point to a less automatic postural control strategy, albeit the correlation between EDSS and balance scores was only significant for EDSS *>* 3^78^.

### EMG

Similar to the collection of IMU data for MS monitoring, walking tasks have been used to collect EMG^72, 79–83^. Furthermore, popular were contraction and extensions tasks^84–87^ as well as reaching tasks^88, 89^. EMG sensors have also been used to investigate dysphagia^90–92^. It is known and apparent that MS induces alterations in muscle synergies, leading to different activation patterns compared to healthy controls. Authors in^79–81^ postulate that these differences may represent compensatory strategies employed by PwMS to cope with motor impairments. Moreover, increased muscle coactivation is observed in PwMS, reflecting an abnormal pattern of muscle activation during walking^72, 79, 82^. Rehabilitation interventions have shown promising results in reducing inappropriate muscle coactivation, thereby potentially improving walking patterns in individuals with MS^72^. Furthermore, after rehabilitation, activation indexes of muscle modules become more similar between people with MS and healthy controls, indicating a potential positive impact of rehabilitation on muscle function^83^. Aside from that, previous electromyographic studies of swallowing in individuals with MS have revealed increased electrical activity in the masseter and decreased activity in the suprahyoid muscles^90^. These changes have been correlated with the Expanded Disability Status Scale (EDSS), suggesting their potential as clinical markers of disease severity. Additionally, people with MS exhibit longer swallowing durations and employ more compensatory respiratory cycles during swallowing^91^. Abnormalities in swallowing are prevalent among individuals with MS, with 92% of patients presenting at least one anomaly during swallowing^92^. Notably, the duration of suprahyoid/submental muscle activity during swallowing has been found to correlate with clinical measures^92^. Studies on reaching tasks have also shown that individuals with MS experience a decrease in modularity and timing delay between mild, moderate, and severe ambulant cases^88^. This implies altered muscle coordination during reaching movements, potentially contributing to motor impairments. In contrast to other research findings,^89^ reported no significant correlation between median frequency in EMG data and subjective fatigue in PwMS and HC after performing shoulder anteflexion in between the goal-directed movements. During contractions and extensions, individuals with MS exhibit lower relative decreases in torque during repeated maximal voluntary contractions, indicating reduced muscle fatigability^84^. However, no significant changes in muscle coactivation were found^84^. Force rate development has been identified as an essential correlate of disability status, possibly outweighing maximal muscle contraction^85^. Rehabilitation interventions have shown potential for enhancing maximal neural drive during knee extensions, as indicated by integrated EMG measures^86^. However, no significant changes were observed in mechanical and electrical fatigue after two weeks of rehabilitation, despite higher force output^87^. Table 1 summarises the main clinical findings when using time-series biosignals, namely EEG, IMU, and EMG data.

**Table 1.** Selection and aggregation of clinical findings in various biosignals.

| Domain | Task | Clinical Findings |
| --- | --- | --- |
| EEG and EPs | VEP<sup>7–37</sup>, MEP<sup>14, 20, 21, 25, 31, 32, 38–45</sup>,<br>AEP<sup>8, 16, 17, 28</sup>,<br>SSEP<sup>10, 17, 20, 21, 28, 35, 36, 43, 46–48</sup> | Longer VEP latencies correlate with retinal fibres are below norm<sup>7, 15, 18, 23, 27, 37</sup><br>Combination of EPs can predict MS progression after 20 years<sup>11</sup>, falls within one year, EDSS longitudinally<sup>20, 21, 25, 28, 31, 46</sup>, brain stem dysfunction<sup>17</sup><br>Combination of EPs show stronger correlation with baseline EDSS than MRI white lesion burden<sup>28</sup> |
|  | Oddball paradigm<sup>49, 63</sup> | The delayed P300 peak reveals the inverse correlation between its amplitude and the mental fatigue level<sup>49</sup><br>ERPs during the visual oddball task are correlated with processing speed at baseline in PwMS<sup>63</sup> |
|  | Resting state<sup>52, 53, 55, 57, 58, 60, 65</sup> | High theta\beta ratio is correlated with slow SDMT speed<sup>60</sup><br>PwMS have lower posterior dominant rhythm and amplitude than HC. This is negatively correlated with disability<sup>65</sup><br>A pronounced correlation between monitoring parameters and annual relapse rate, disability score, depression score and level of fatigue<sup>52</sup> |
| IMU | Walking<sup>67–73</sup> | Significant correlation between gait parameters (normalized velocity, stride length<sup>67, 73</sup> and hip range motion<sup>67</sup>, higher variability in their lumbar yaw range and lateral foot deviation<sup>68</sup>, asymmetrical sway patterns<sup>69</sup>, stride regularity and swing time asymmetry<sup>70, 73</sup>). |
|  | Sit Stand Transitions<sup>75, 75–77</sup> | Accelerometer-based metrics can separate fallers from non-fallers<sup>75, 76</sup><br>Correlation between accelerometer-based performance metrics and MS disease severity, fatigue and balance confidence<sup>77</sup> |
| EMG | Walking<sup>72, 79–83</sup> | Muscle co-activation differs between PwMS and HC<sup>72, 79, 82</sup> which may represent compensatory strategies to cope with motor impairments |
|  | Contraction<sup>84–87, 89</sup> | Lower relative decreases in torque during repeated maximal voluntary contractions<sup>84</sup><br>Force rate development has been identified to correlate with disability status<sup>85</sup> |
|  | Swallowing<sup>90–92</sup> | Increased swallow durations and more compensatory respiratory cycles<sup>91</sup><br>92% of patients presenting at least one anomaly during swallowing<sup>92</sup><br>Differences in electrical activity in the masseter and in the suprahyoid muscles<sup>90</sup> |

### Speech

In the scope of multiple sclerosis (MS) research (monitoring and diagnostics), speech analysis has recently emerged as a promising approach for monitoring the progression of the disease. The reviewed literature explored various speech tasks to examine changes in speech production in PwMS. Several studies have focused on the task of sustaining the vowel /a/ for extended durations^93–97^, as well as the diadochokinetic syllable repetitions of /pa/-/ta/-/ka/^93, 94, 97, 98^. Furthermore, investigations have included tasks such as naming the days of the week^93, 97^, reading tasks^93, 94, 97–102^, monologue engagements^98^, narrating stories with positive content from memory^93, 97^, and describing pictures or individuals^100, 103^. Additionally, the exploration of tasks involving the production of the vowel /a/ at the highest pitch, phonation of the vowel /a/ at the lowest possible volume and reading words with a consonant-vowel-consonant (CVC) syllable structure has also been documented^95^. Dual-task speech performance in MS patients^103^ and expiratory rate in MS patients^96^ were also explored. Combining multiple speech measures was found to facilitate the prediction of cerebellar impairment in PwMS^93^, and machine learning models combined with automatic voice recording analysis showed promise in diagnosing the disease^99^. Notably, speech analysis from reading tasks exhibited a high accuracy in distinguishing between PwMS and HC^99^. Speech features also demonstrated strong correlations with clinical markers and commonly used tests in MS evaluations, such as 9HPT, EDSS, T25FW and PASAT^101^. Furthermore, comparisons between PwMS subgroups and HC highlighted several key findings. Patients with PPMS exhibited reduced articulation and speech rates compared to HC and patients with RRMS^100^. These changes in speech rate correlated with bilateral white and grey matter loss, brain atrophy, and the EDSS. Similarly, diadochokinetic rate correlated with brain volume changes, EDSS score and MSFC^94^. Regarding speech patterns in PwMS, cognitive impairment was also examined. Reduced cognitive resources resulted in specific patterns in reading behaviours, leading to slower articulation and speech rates during reading tasks. Prosodic features and other speech and language characteristics showed promise as potential markers for evaluating cognitive impairment in MS^102^. The correlation between cognitive measures and articulation rate further supports using objective speech analysis in identifying patients with cognitive impairment^98^. Patients were successfully classified into three subgroups of EDSS (mild, moderate, severe) using an acoustic composite score that showed correlations with EDSS, multiple sclerosis impact scale (MSIS), total lesion load, and white matter volume^97^. Expiratory time demonstrated significant correlations with EDSS scores. MS patients exhibited lower maximum phonation times and maximum expiratory times compared to healthy controls^96^. Additionally, dysarthria significantly affected speech timing patterns, resulting in slower speech with longer pauses. Patients with cognitive impairment and dysarthria faced difficulty maintaining speech timing patterns, highlighting the complex interactions between dysarthria and cognitive impairment^104^. It is worth noting that dual tasks significantly affected speech rate and sentence duration in both PwMS and HC, with total sentence duration differing significantly between the two groups^103^. Additionally, measures such as formant centralization ratio (FCR) and dysphonia severity index (DSI) score were significantly different between HC and PwMS. Additionally FCR correlated with EDSS and disease duration^95^. Table 2 summarises the previously investigated speech tasks, extracted acoustic features, and the main clinical outcomes and findings of these clinical studies.

**Table 2.** Summary of the different speech studies on MS and their main clinical findings.

| Speech Task | Features | Clinical Findings |
| --- | --- | --- |
| Sustaining a vowel /a/<sup>93-97,101</sup> | Frequency Instability (Jitter), Loudness Instability (Shimmer), Harmonics-To-Noise Ratio, Cepstral Peak Prominence, Maximum Phonation Time | Used in composite score to predict cerebellar dysfunction in MS<sup>93</sup><br>Combined with articulatory decay distinguishes between HC and MS (77.5% accuracy)<sup>101</sup><br>Used in DSI which differs significantly between HC and PwMS<sup>95</sup><br>PwMS have lower max phonation time<sup>96</sup> |
| Diadochokinetic Speech<sup>93,94,97,98,101</sup> | Diadochokinetic Speech Rate (syllables/second), Articulatory variability (syllables length SD), Diadochokinetic Regularity | Used in composite score to predict cerebellar dysfunction in MS<sup>93</sup><br>Diadochokinetic speech rate was correlated with cerebellar grey and white matter fraction and brain parenchymal fraction<sup>94</sup><br>Part of the acoustic composite score which separated PwMS into three EDSS subgroups<sup>97</sup> |
| Reading<sup>93-95,97-102</sup> | Speech Duration, Speech Rate/Tempo, Pause Characteristics, Articulation and Vowel Characteristics, Intensity and Loudness Measures, Spectral Characteristics | Can predict health status (82% accuracy)<sup>99</sup><br>Patients with PPMS have a reduced articulation and speech rate compared to HC and patients with RRMS<sup>100</sup><br>Articulation rate is correlated with bilateral white and grey matter loss, brain atrophy and EDSS<sup>94</sup><br>Used in the Formant Centralization Ratio that correlates with EDSS and disease duration<sup>95</sup><br>Reduced cognitive resources can lead to slower articulation/speech rate during reading tasks<sup>102</sup> |
| Free Speech<sup>93,97,98,100,103,104</sup> and Automatic Speech (naming days of the week)<sup>93,97</sup> | Speech Duration, Speech Rate/Tempo Characteristics, Pause Characteristics, Spectral Characteristics | Used in composite score to predict cerebellar dysfunction in MS<sup>93</sup><br>Patients with PPMS have a reduced articulation and speech rate compared to HC and patients with RRMS<sup>100</sup><br>Dysarthria reflects in slower speech with longer pauses in PwMS and cognitive impairment interactions<sup>103</sup> |
| Phonation of vowel /a/ (highest freq. and lowest intensity)<sup>95</sup> | Maximum F0, Minimum Intensity | Used in DSI which differs significantly between HC and PwMS<sup>95</sup> |
| Dual Task<sup>103</sup> | Speech Rate, Articulation Rate, Silent Pause Duration, Silent Pause Frequency, Sentence Duration, Onset Reaction Time | The total sentence duration in the dual-task was significantly different between PwMS and HC<sup>103</sup> |

### Sensor-Free Digital Apps

Over the last few years, we have witnessed a huge interest in using sensor-free digital apps to monitor PwMS. Such digital apps have sparked the interest of pharmaceutical companies, whose focus is on monitoring treatment response and disease progression. Hence, such apps help them personalize and tailor their treatments based on the patient’s needs, phenotype, and disease stage. In this context, several mobile apps have been developed for remote monitoring multiple sclerosis^105–108^. The apps provide a set of tasks to evaluate different function systems affected by the disease. For cognition, apps used digital symbol substitution tests^105, 106^. Dexterity was covered with pinching and drawing shape tasks^105^, finger-tapping^106^ and a screen-to-nose test^108^. Balance and walking were rated with timed distance walking^105–108^ and sit-stand transitions^108^. Tasks were correlated with the results of their clinical counterparts, such for example, the SDMT for cognition^105^ and 9HPT for dexterity^108^. The results of combined scores were successfully correlated with corresponding established clinical scores, such as the EDSS^105, 107, 108^ and MSFC^107^.

### Other Wearables

During a typing analysis with a custom on-screen smartphone keyboard, the MS group had slower typing speed, which was also associated with processing speeds.^109^ Furthermore, PwMS with better cognitive function showed a higher correlation between backspace and autocorrection events.^109^. Hilty et al. found an association between heart rate variability and disease severity in PwMS through 24-hour monitoring with a wearable device^110^. During a virtual peg insertion test, smoothness and grip force measured with a haptic device were altered in PwMS compared to HC and showed higher sensitivity than its clinical counterparts^111^. A wearable device’s passive monitoring of features, including physical activity duration, step count, active energy expenditure, and metabolic equivalents, successfully computed a composite score that correlated with brain volume loss^112^.

## Discussion

### Remote monitoring could reduce clinical and economic MS cost burden and enable personalised MS treatments

This paper systematically reviewed the world’s last decade’s literature regarding digital health and the use of biosignals for MS monitoring and management (both home & hospital-based monitoring). Noticeably, most of the reviewed papers solely focused on analysing patients at a particular time interval and did not include the disease’s evolution aspect nor clinically study or monitor patients longitudinally. Nonetheless, a few studies managed to track the same patients over time and evaluated MS progression. While we initially wanted to focus on remote monitoring, the findings from this review highlight that only a few studies were conducted in an unsupervised home environment and remote fashion, which might be due to usability, patient compliance and adherence, or privacy concerns. To exemplify this, studies including speech for remote MS monitoring were neither longitudinal nor unsupervised. Conversely, such an approach has successfully been tested with other akin brain disorders, such as Parkinson’s^113^. This comprehensive systematic review confirms a gap in the literature when using digital health, biosignals and remote monitoring technology to monitor MS patients. These identified knowledge gaps have had implications for moving clinical evidence into practice and bringing it to patient’s hands. Hence, such gaps contribute to making MS, according to a recent US study^114^, the second most expensive chronic disease after heart failure, leading to enormous economic and clinical cost burdens. Consequently, introducing and adopting remote monitoring technology and digital health would have significantly reduced these burdens and costs by enabling the right patient to receive the proper treatment at the right time and under the right circumstances. Unquestionably, clinical outcomes, such as reduced disability, reduced relapse rates, better workplace productivity, higher life expectancy, and improved quality of life, could be achieved thanks to remote monitoring and management technologies^114^. Interestingly, clinically assessing the available MS treatment options based on monitoring technology has not been extensively investigated. While various clinical attempts used biosignals and monitoring technologies and studied the correlation between their findings and patients’ scores of disability or even predicted MS progression, this review highlights that studying how to personalise and individualise treatment (frequency, duration, and treatment response) has rarely been investigated^115^. Remarkably, a recent (2022) study by a pharmaceutical company (Merck) explored the use of machine learning and digital health monitoring technology to determine the optimal treatment duration for MS patients on Mavenclad medication^115^. Their promising findings should hopefully spur further research on the use and adoption of similar monitoring technology to assess treatment response and effectiveness, as well as investigate how to personalise it based on the patient’s phenotype and disease course.

### Barriers to Remote Monitoring Adoption: Identified Gaps & Technical Challenges

Throughout this systematic literature review, it has become apparent that different technical challenges and gaps exist in remote MS monitoring and management. Our review’s findings add a rehabilitation dimension to the literature concerning the lack of advanced technological solutions for remote MS patient monitoring. With the exception being EPs and tasks inside the mobile applications using MSFC^105–108^, to the best of our knowledge, there have been no attempts to expand the feature sets of predicting disability and progression across multiple domains. For instance, speech is almost entirely evaluated isolated, without considering other digital biomarkers, such as EPs from EEG. Since the combination of EPs (known as multi-modal EPs) has already shown more accurate results over the use of individual EPs, it is thought that the combination of multiple digital biomarkers (biosignals) would further improve the prediction of disability and progression. The popularity and momentum machine learning has gained over the last few years^116^ has spurred its use in MS patient monitoring. The findings of this review accentuate that the most frequently used machine-learning models in previous studies are linear and paired with a statistical analysis of correlations of single parameters. While this approach makes it easy to get an intuition about the relations, it is worth exploring more advanced machine learning models to boost predictive power and explore more complex features and digital biomarkers. Aside from that, the impact of MS on the latency of EPs is widely understood, but reducing the resolution of the signal to a single measure and thus losing information that is entangled, for example, within the shape of the movement, might be a missed opportunity. This could be observed in the work of Yperman et al., who found that the latency of MEPs is not the most predictive feature when incorporating other time-series features^41^. More advanced models do not necessarily come at the cost of explainability, as shown by the treatment duration optimization of Mavenclad^115^. The findings of this review point out the need for higher-resolution wearable sensors whose sensitivity and accuracy are suitable for use in a noisy home (patient) environment. Whilst the MSFC already shows greater sensitivity compared to the EDSS^117^ (particularly for patients with lower disabilities), including more powerful sensors with higher resolution could allow for even higher sensitivity. Tasks, such as the T25FW, for instance, do not evaluate gait patterns in terms of variability but only consider the overall velocity. Over and above, SDMT for evaluating cognition can be influenced by practice effects^118^, making it the only source of cognitive information, which may not be sufficient. As demonstrated by some papers^53, 63^, there are other ways to assess cognitive function, which are less vulnerable to practice effects^53, 63^.

### Data Privacy and Security Issues: Gaps & Challenges

Unequivocally, the widespread use of wearable data and digital health apps in remote MS monitoring and management raises various concerns about data privacy and management. The most pressing questions are orbiting around how we can protect sensitive wearable data from undesired data breaches. Alarmingly, the recent increase in cyberattacks presents a significant issue when using digital health in remote patient monitoring. Another pressing question is how to enable multi-site and decentralised clinical trials in MS, as well as to facilitate wearable data sharing for research and clinical purposes. Novel concepts, such as federated learning (FL)^119^ depicted in Figure 2, could play a paramount role in unlocking that potential and fostering collaborative and decentralised clinical work on MS. With the recent unprecedented interest in using machine learning and artificial intelligence in digital health, new acts and laws, such as the new EU AI Act whose purpose is to regulate the use of AI, has become a necessity. Aside from that, user agreements and e-consents about data transmission frequency, access, and data processing should transparently be discussed and designed between patients and their medical providers.

**Figure 2.**
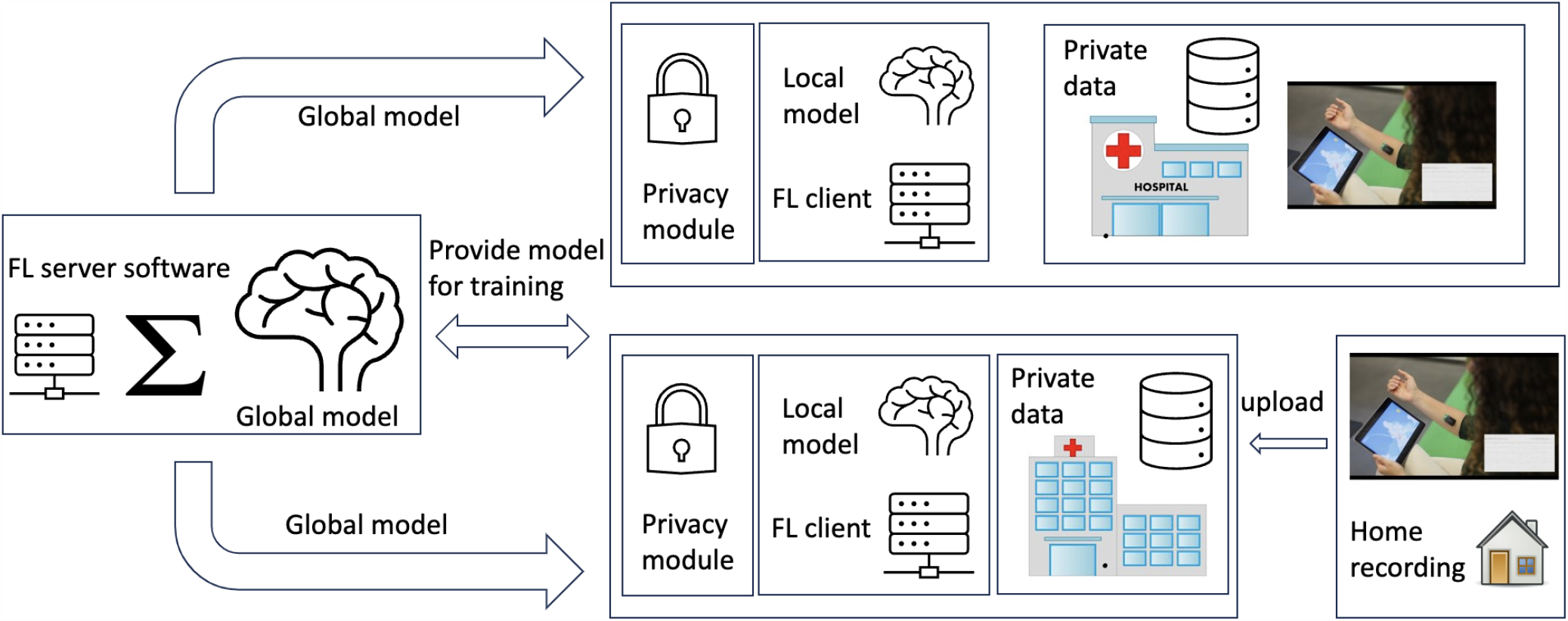
An overview of the Federated learning concept and its potential use in remote MS monitoring

### Future Opportunities

Notwithstanding the highlighted challenges and identified barriers to embracing the power of digital health technology for remote patient MS monitoring, many opportunities lie. Unlike any other brain disorder, MS is commonly diagnosed in the 20s & 30s^2^, making its remote monitoring vitally important. Overall, the total lifetime cost per patient with MS is estimated to be $ 4.1 million, and the total estimated economic burden is around $85.4 billion. Furthermore, the financial and clinical costs increase from $30k to $100k per patient per year when an MS progresses from a relapsing-remitting stage to a more progressive type of MS^120^. It is therefore thought that digital technologies and remote patient monitoring solutions could play an essential role in slowing this progression and thereby avoiding disability. With approximately 39% of MS patients publicly insured and 53% privately covered^120^, payers and insurers in the USA and around the world should consider new reimbursement models and schemes for remote MS monitoring and management. In this post-COVID-19 era, insurers have become more open to accepting and reimbursing remote monitoring solutions for acute and chronic conditions^121^. Thus, reimbursement of software-as-a-medical devices and digital app technologies has gained momentum, and many new reimbursement schemes have been designed and implemented. Similarly, various governmental endeavours and new reimbursement vehicles, such as the Digital health applications (DiGA) system in Germany, PECAN in France, and the breakthrough device designation program by the U.S. Food and Drug Administration (FDA), have recently been implemented and tested. Hence, such policies and reimbursement endeavours will pave the way for bringing clinical research into the MS patient’s home and charting a route ahead for a new era of remote MS patient management and monitoring. From a technological standpoint, we have witnessed the rapid spread of digital health technology and the remarkable interest in adopting novel and groundbreaking technology, such as non-invasive brain-computer interfaces (BCIs)^122^. Non-invasive BCIs, particularly with the newly designed EEG, EMG, and ECG wearable sensors, are expected to be combined with telehealth platforms, leading to a real revolution in MS patient care^123^. Irrefutably, unlocking the full potential of that requires multidisciplinary collaboration between different stakeholders involved in MS care, including payers, insurers, governments, clinicians, and, more importantly, patients themselves.

## Conclusion

In conclusion, this review of more than 100 published studies highlights the gaps, challenges, and barriers to using digital health solutions and wearable sensors in MS monitoring and management. Previous systematic reviews tend to focus on one single aspect of the disease, such as assessing one particular symptom or one single modality (biosignal), or they concentrate on mobile apps using questionnaires, excluding, therefore, the myriad of previously conducted clinical studies using biosignals and different potential digital biomarkers. This review is unique and comprehensive, encompassing over 100 MS studies and covering previous clinical studies in both home environments and clinical settings. Whilst we only included studies in the English language and did not evaluate or examine publication bias, this review’s findings and main drawn conclusions should enhance the reader’s understanding of the gaps in the literature and the obstacles to fully adopting digital health technologies and wearable sensors to monitor MS progression and its treatment effectiveness. It is evident that digital health technologies offer considerable promise and hold an enormous potential to revolutionise the remote monitoring of MS progression and assessment of patient treatment effectiveness and response. With high treatment costs and the lack of a one-size-fits-all approach in MS treatment prescription, this review concludes that personalised medicine and remote monitoring solutions are crucial to slowing down MS progression, improving treatment response, and avoiding the devastating outcome of disability. In the post-COVID-19 era, remote and decentralised patient care has sparked interest and spurred research in that direction. This review foresees current innovations in digital health technology for MS monitoring and management to be catalysed and fully adopted in the near future. This will, therefore, open up limitless opportunities to combat silent MS disability progression and significantly improve patients’ quality of life.

## Data Availability

No data was collected as the paper is a systematic review.

## Additional information

### Author contributions

All authors contributed to this research. B.S. performed the research, conducted the literature review and wrote the paper. H.J. conducted the literature review of the speech part. S.G. assisted with reviewing the EEG literature review and summarizing that part. A.P., D.K., P.A.S, R.S., and R.C. co-supervised the work and reviewed the paper. Z.T. designed the research, supervised all the work, and co-wrote the paper. All authors contributed to writing the paper.

## Acknowledgement

The authors would like to thank the Ministry of the Economy in Luxembourg and its digital health directorate for supporting this research. Similarly, we would like to thank the Luxembourg National Research Fund (FNR) for funding this research.

